# JIA patient T cells differentiate into Th1, Th17 and Th1.17 effector cells under Th1 polarizing conditions

**DOI:** 10.1101/2021.10.01.21264425

**Authors:** Anna E. Patrick, Tashawna Esmond, Kayla Shoaff, David M. Patrick, David K. Flaherty, T. Brent Graham, Philip S. Crooke, Susan Thompson, Thomas M. Aune

## Abstract

**Objective:** T helper cells develop into discrete Th1, Th2 or Th17 lineages that selectively express IFNγ, IL-4/IL-5/IL-13, or IL-17, respectively and actively silence signature cytokines expressed by opposing lineages. Our objective was to compare Th1, Th2 and Th17 polarization in cell culture models using JIA patient samples.

**Methods:** Peripheral blood mononuclear cells were isolated from JIA or healthy prepubescent children. T cell naïve and memory phenotypes were assessed by flow cytometry. T cell proliferation was measured using a fluorescence-based assay. Th cell cultures were generated *in vitro* and IFNγ, IL-17, and TNFα measured by ELISA and flow cytometry.

**Results:** JIA Th1 cells produced increased IFNγ and inappropriately produced IL-17. JIA Th17 cells produced increased IL-17. JIA Th1 cell cultures develop dual producers of IFNγ and IL-17, which are Th1.17 cells. JIA Th1 cultures expressed elevated levels of both T-bet and RORγT. RNA sequencing confirmed activation of immune responses and inappropriate activation of IL-17 signaling pathways in Th1 cultures. A subset of JIA patient samples was disproportionally responsible for the enhanced IFNγ and IL-17 phenotype and Th1.17 phenotype.

**Conclusions:** This study reveals that JIA patient uncommitted T cell precursors, but not healthy children, inappropriately develop into inflammatory effector Th1.17 and Th17 cells under Th1 polarizing conditions.

**Rheumatology key messages:**

- Th1 differentiation of JIA PBMCs generates high IFNγ, IL-17, and dual IFNγ-IL-17 producing cells.
- JIA Th1 differentiation increases master transcription factor expression for Tbet and RORγT.
- Enhanced JIA Th1 IFNγ and IL-17 production occurs in a subset of JIA patients.

## Introduction

Juvenile idiopathic arthritis (JIA) is the most common autoimmune arthritis in children. Effective therapies for JIA include disease-modifying anti-rheumatic drugs (DMARDs) and biologics that target T cell activation and inflammatory cytokines (1, 2). Most JIA patients attain inactive disease with therapies; however, the likelihood of disease flare is over 50% in the first 2 years after attaining inactive disease (3, 4). Moreover, chronically uncontrolled JIA occurs in half of patients that require multiple DMARDs during treatment (5). Critical needs exist for a personalized approach to current therapies and development of new therapies. Meeting this critical need is hindered by our poor understanding of how inflammatory T cells and cytokines develop in JIA.

In JIA, the clinical and biologic phenotypes are diverse (6). JIA patients with the same clinical subtype can have different biologic phenotypes. Clinical subtypes include more or less involved joints at onset (polyarticular and oligoarticular respectively) and are further subdivided by criteria such as rheumatoid factor (RF), and HLA-B27 positivity and presence of psoriasis in patient or family members (7). The systemic JIA subtype has features of autoinflammatory disease and is distinct (7). The peak JIA incidence is 1-5 years old (8), which is especially true for oligoarticular and polyarticular RF negative subtypes. At this age, the immune system is shifting from more naïve to memory T cells (9, 10). In JIA conflicting evidence exists about whether naïve and memory T cell frequencies are different from healthy children (11-13).

T cells include CD4^+^ T helper (Th) cells that coordinate immune responses by producing cytokines to stimulate the immune system to combat infections. Th1, Th2, and Th17 cells differentiate from naïve CD4+ cells in response to T cell receptor stimulation and combinations of polarizing cytokines. These cells selectively express signature cytokines: Th1: IFNγ and TNFα, Th2: IL-4, IL-5, and IL-13, TH17: IL-17 (14-17). Key transcription factors are critical for differentiation of each Th lineage: Th1: Tbet, Th2: GATA3, Th17: RORγT and BATF. During differentiation, each pathway suppresses alternate pathways. Th1 and Th17 cells and their effector cytokines are inflammatory. In polyarticular and oligoarticular JIA, more Th1 and Th17 cells are present in the synovial fluid of joints with arthritis (18-21). Furthermore, a Th1-like Th17 cell, Th17.1, is present in synovial fluid and considered pathologic (22). The origins of these cells are unclear.

We find that JIA peripheral blood mononuclear cells (PBMCs) undergo abnormal Th cell differentiation and inappropriately produce inflammatory cytokines, IFNγ and IL-17, in well-established short-term T cell cultures that model Th cell differentiation and cytokine production (23, 24). With a focus on prepubescent children, we hypothesize that JIA precursor Th cells are predisposed to heightened inflammation under Th cell polarizing conditions.

## Methods

### Patients and samples

JIA PBMCs from Vanderbilt Monroe Children’s Hospital and the Cincinnati Children’s Hospital Pediatric Rheumatology Tissue Repository, HC^C^ PBMCs from the Cincinnati Children’s Hospital Medical Center genomic control cohort, and HC^A^ PBMCs from Vanderbilt were all obtained with IRB approved protocols. Isolated PBMC were stored in fetal bovine serum with 10% DMSO in liquid nitrogen until use.

### Th cell cultures and cytokines

PBMCs were cultured under Th1, Th2, and Th17 differentiation conditions in a well-defined tissue culture model for Th cell differentiation and cytokine secretion (23, 24). Cultured cells were in RPMI 1640 (Gibco) supplemented with 10% fetal bovine serum (Atlanta Biologicals), L-glutamine (Gibco), and penicillin-streptomycin (Gibco) with 5% air CO_2_. PBMCs with 100,000 cells/well in 96-well plate or 500,000 cells/well in 24-well plate were stimulated on anti-CD3 (10µg/mL) precoated plates with addition of anti-CD28 (1µg/mL) and stimuli for Th cell differentiation. Stimuli are Th1: IL-12 (10ng/mL), Th2: IL-4 (10ng/mL), Th17: IL-6 (50ng/mL), IL-1β (20ng/mL), TGF-β1 (2ng/mL), IL-23 (20ng/mL), IL-21 (100ng/mL), anti-IFN*γ* (10µg/mL). Purchased reagents are antibodies from Biolegend and cytokines from BD Biosciences and R&D Systems. After 5 days for Th1 and Th2 cultures or 7 days for Th17 cultures, cells were washed with complete media and re-stimulated on new anti-CD3 coated plates. After 2 days, culture fluids and cells were harvested.

Cytokines were analyzed by ELISA to detect human IFNγ (BD Biosciences), TNFα (BD Biosciences), IL-17a (ThermoFisher), IL-5 (BD Biosciences), and IL-13 (ThermoFisher). Th cultures were performed in triplicate and averaged.

### Flow cytometry

PBMC or Th cells were analyzed with LIVE/DEAD^™^ Fixable Violet Dead stain (Life Technologies), anti-CD3 APC-H7, anti-CD8a PE, anti-CD4 perCP-cy5.5, anti-CD197 AF647, and anti-CD45RA FITC using standard flow cytometry protocols. Gating was performed with fluorescent minus one (FMO) control for CD197^+^ and CD45RA^+^ populations. Experiments were performed on a 3-Laser BD LSRFortessa instrument maintained by the Vanderbilt University Medical Center Flow Cytometry Shared Resource. Th1 cells were analyzed from Th1 cultures that had been restimulated on day 5 and treated with PMA (30ng/mL), ionomycin (1µg/mL), and GolgiStop (1µL/mL) for 6 hours on day 7. Cells were analyzed with Fixable Zombie NIR, anti-IFNγ PEDazzle594, anti-IL-17 APC, and anti-Tbet PE-Cy7 using flow cytometry standard protocols. Gating was performed with FMO control for IFNγ^+^ and IL-17^+^ populations. Experiments were performed on a Cytek Aurora instrument. Purchased reagents are antibodies from BD Pharmingen and Biolegend and reagents from BD Biosciences. Analysis performed using FlowJo software.

### Cell proliferation

PBMCs labeled with carboxyfluorescein succinimidyl ester (CFSE)(10µM) using the CellTrace CFSE Cell Proliferation Kit (ThermoFisher, C34554) were stimulated with immobilized anti-CD3 and anti-CD28 (1µg/mL) and in some experiments IL-12 (10ng/mL). On day 3, harvested cells were analyzed by flow cytometry for live/dead cells, anti-CD3 APC-H7, anti-CD8a BV711, anti-CD4 APC. Purchased reagents are from Life Technologies, BD Pharmingen, BD Biosciences, and Biolegend. Experiments were performed on a 3-Laser BD LSRFortessa instrument. Analysis was performed using FlowJo software Proliferation Modeling.

### RNA analysis

RNA was collected from Th1 and Th2 cell cultures differentiated for 5 days using TRIreagent and complementary DNA synthesized using SuperScript First Strand Synthesis (ThermoFisher). Quantitative real time PCR measured expressed RNAs using SYBR green master mix (Applied Biosystems). RNA levels were standardized relative to the housekeeping gene GAPDH. Definite outliers were removed using ROUT with Q=0.1% by GraphPad Prism software.

For RNA sequencing, RNA was collected by TRIreagent followed by DNase digestion. Library preparation with the Illumina Tru-Seq Stranded RNA kit and RNA sequencing with an Illumina HiSeq2500 instrument were performed by the Vanderbilt Technologies for Advanced Genomics (VANTAGE) core facilities consecutively on all samples. 100bp paired end reads were generated. Average sequencing depth of all samples was 43 million mapped reads ± 13.5 million (standard deviation). FASTQ files were processed using DESeq2 to identify differentially expressed genes (26). GO Enrichment analysis for overrepresented biological processes was performed using the Gene Ontogeny Resource (27-29).

### Statistical analysis

Applied statistical tests were Mann-Whitney test for comparison of 2 and Anova for comparison of more than 2 samples. P < 0.05 was considered significant.

## Results

### Clinical characteristics of JIAs and healthy controls

We performed studies in prepubescent children with JIA and age-, gender-, and race-matched controls (HC^C^) (Table 1). JIA patients were an average of 62 months old, ANA+, and mostly female, consistent with known clinical characteristics. JIA subtypes were defined using International League Against Rheumatism (ILAR) criteria (25). Clinical characteristics were collected at the time of PBMC collection (Table 1). Five JIA PBMCs were collected within 1 month of diagnosis and before initiation of systemic medications other than non-steroidal anti-inflammatories (NSAIDs). This group had more active joints with arthritis. Overall, JIA patients had 0-1 active joints with arthritis and were on methotrexate and/or biologic medications including etanercept, adalimumab, infliximab, and abatacept.

**Table 1.** Demographic and clinical characteristics of JIA or healthy control groups.

|  | HC<sup>c</sup> | JIA |
| --- | --- | --- |
| Number of cases | 20 | 20 |
| Age in months (range) | 62 (36-93) | 62 (24-94) |
| Gender, female, n (%) | 16 (80%) | 16 (80%) |
| Ethnicity, Caucasian, n (%) | 19 (95%) | 19 (95%) |
| ANA positive, n (%)<sup>*</sup> |  | 19 (95%) |
| JIA subtypes, n (%) |  |  |
| Polyarticular RF- |  | 17 (85%) |
| Polyarticular RF+ |  | 1 (5%) |
| Oligoarticular |  | 2 (10%) |
| **<u>Characteristics at Time of PBMC Collection</u>** |  |  |
| Active joints, n (%) |  |  |
| 0-1 |  | 12 (60%) |
| 2-4 |  | 3 (15%) |
| >4 |  | 5 (25%) |
| Disease duration <1 month, n (%) |  | 5 (25%) |
| Time since diagnosis in months (range) |  | 17.5 (0-70) |
| Physician global assessment score (range) |  | 2 (0-9) |
| Current medications, n (%) |  |  |
| Methotrexate only |  | 5 (25%) |
| Biologic only |  | 3 (15%) |
| Methotrexate + Biologic |  | 6 (30%) |
| None or NSAID |  | 5 (25%) |

<sup>\*</sup>1 ANA unknown, child healthy controls (HC<sup>c</sup>)

### Frequencies of naïve and memory T cells in JIA, HC^C^, and HC^A^ and T cell proliferation

Differences between JIA and HC^C^ peripheral blood mononuclear cells (PBMCs) were investigated by analyzing T cell subsets and naïve and memory phenotypes. JIA, HC^C^, and adult control (HC^A^) PBMCs had the same frequencies of CD3^+^, CD3^+^CD4^+^, and CD3^+^CD8^+^ cells (Supplemental Figure 1). On average, 60% were CD3^+^ cells, of which approximately 60% were CD4^+^ and 30% were CD8^+^.

Memory and naïve cell phenotypes were identified by expression of CD197 and CD45RA with naïve cells: CD197^+^CD45RA^+^, central memory cells: CD197^+^CD45RA^-^, and effector memory cells: CD197^-^CD45RA^-^. Pediatric (JIA and HC^C^) CD3^+^CD4^+^ cells were an average of 69% naïve and 23% combined central and effector memory cells. This is opposed to adult CD3^+^CD4^+^ cells that were an average of 44% naïve and 54% combined central and effector memory cells (Figure 1A,B). Pediatric CD3^+^CD8^+^ cells were an average of 70% naïve and 12% combined effector and memory cells. This is compared to adult CD3^+^CD8^+^ cells that were an average of 52% naïve and 31% combined effector and central memory cells (Figure 1C,D). In pediatric patients, ratios of naïve to memory CD3^+^CD4^+^ and CD3^+^CD8^+^ cells were 3 and 5.8 respectively, while in adults, ratios were reduced at 0.81 and 1.6. Importantly, no differences in memory and naïve cell phenotypes were identified between JIA and HC^C^.

**Fig. 1.**
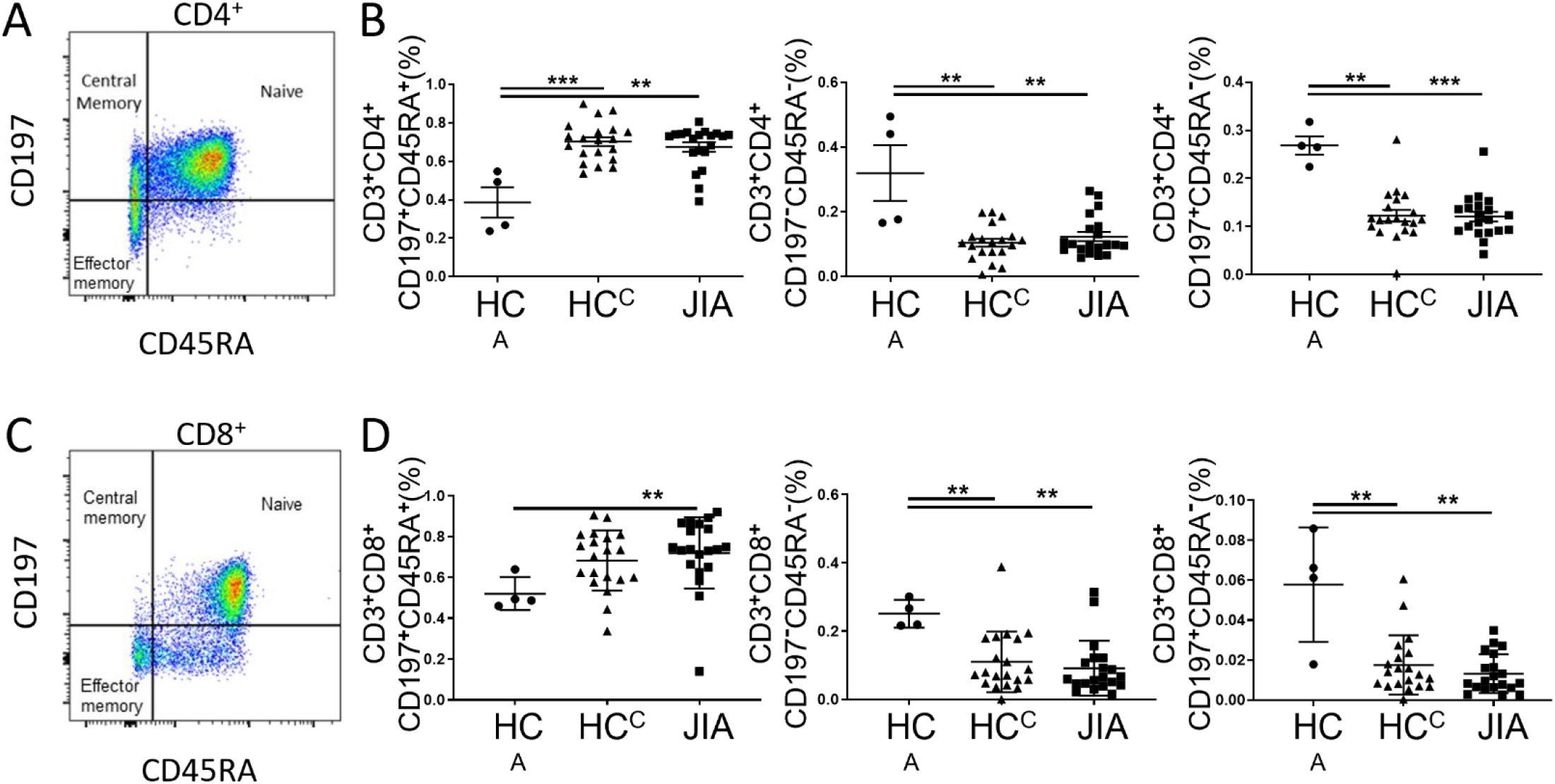
Naïve and memory T cells in adult and child healthy controls and JIA. Adult healthy control (HC^A^), child healthy control (HC^C^), and JIA PBMCs were analyzed with flow cytometry for CD3^+^CD4^+^ and CD3^+^CD8^+^ naïve (CD197^+^CD45RA^+^), central memory (CD197^+^CD45RA^-^), and effector memory (CD197^-^CD45RA^-^) cell subsets. (A) CD3^+^CD4^+^ subtype representative flow cytometry plot from JIA. (B) Analysis of naïve, effector memory, and central memory frequency of CD3^+^CD4^+^ cells. (C) CD3^+^CD8^+^ subtype representative flow cytometry plot from JIA. (D) Analysis of naïve, effector memory, and central memory frequency of CD3^+^CD8^+^ cells. Adult (N=4), JIA (N=20), and child control (N=20). Shown is mean with standard deviation. * P < 0.05, ** P< 0.01, *** P<0.001 by Mann-Whitney test.

JIA and HC^C^ T cell proliferation was assessed by labeling PBMCs with the fluorescent dye, CFSE. Labeled cells were stimulated with anti-CD3 and anti-CD28 and proliferation measured on day 3 (Supplemental Figure 2A). The proliferation index (PI), which is the number of divisions per dividing cell, for JIA and HC^C^ were not different. On average, PIs for CD3^+^, CD3^+^CD4^+^, and CD3^+^CD8^+^ cells were 1.4, 1.4, and 1.5 (Supplemental Figure 2B). The impact of Th1 polarization on T cell proliferation was determined by adding IL-12 to the proliferation assay. Again, we found PIs for JIA and HC^C^ were not different. On average, the PIs for Th1 polarized CD3^+^, CD3^+^CD4^+^, and CD3^+^CD8^+^ cells were 1.8, 1.7, and 1.9 (Supplemental Figure 2C). The PIs for Th1 polarized cells were higher than non-polarized cells when JIA and HC^C^ were combined (Supplemental Figure 2D). Importantly, no differences in PIs were identified between JIA and HC^C^.

### Aberrant cytokine expression by JIA effector Th cell cultures

We determined if peripheral JIA T cells, which were largely naïve, develop inflammatory cytokine profiles with Th cell differentiation. We used short-term Th cell cultures from peripheral blood to model Th cell differentiation and cytokine production (23, 24). JIA and HC^C^ PBMCs underwent Th1, Th2, and Th17 differentiation, reactivation, and the inflammatory cytokines IFNγ, IL-17, and TNFα were measured by enzyme-linked immunosorbent assay (ELISA). In Th1 cultures, JIA exhibited increased production of IFNγ (Figure 2A). Notably, while Th2 cultures should fully suppress IFNγ production, IFNγ was detectable in several JIA Th2 cultures. JIA Th1 and Th17 cultures had increased production of IL-17 (Figure 2B). Notably, IL-17 production in JIA Th1 cultures was similar or even greater than its production in HC^C^ Th17 cultures. No differences were detected in TNFα levels (Figure 2C). In Th2 cultures IL-17 and TNFα production were below the limits of detection. To determine if the 5 newly diagnosed JIA have a skewed pattern inflammatory cytokine production, these patients were identified in our Th culture analysis (Figure 2, red circles). The newly diagnosed JIA were not different than total JIA for Th1, Th2, and Th17 culture production of IFNγ, IL-17, and TNFα.

**Fig. 2.**
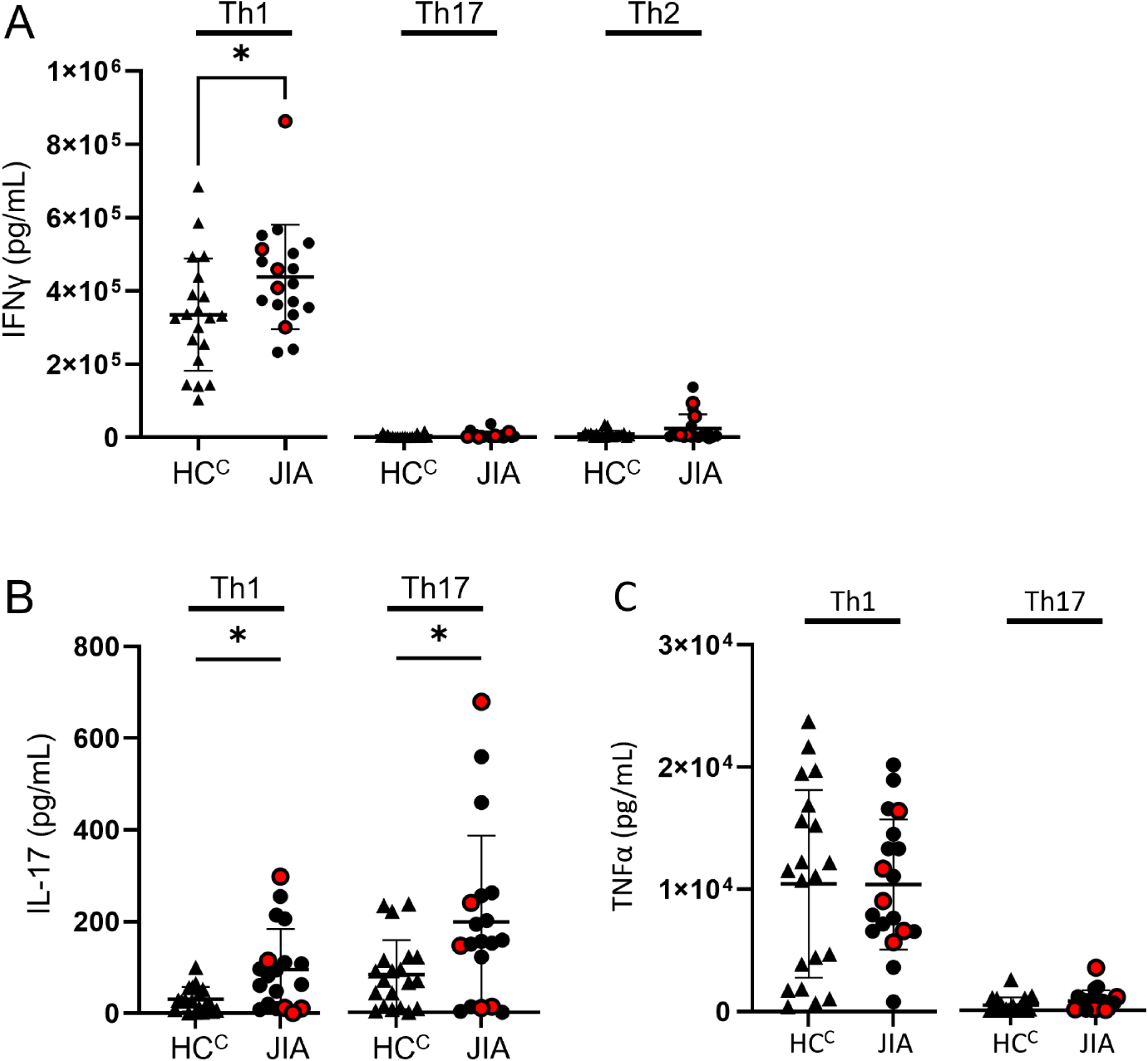
Increased production of IFNγ and IL-17 from JIA T helper cells. Th1, 2, and 17 cell cultures were differentiated *in vitro* from child healthy control (HC^C^) and JIA PBMCs. After for 5 days for Th1 and Th2 or 7 days for Th17 cultures, cells were reactivated, and the produced cytokines measured by ELISA after 2 days. (A) IFNγ produced by Th1, Th17, and Th2 cells for HC^C^ (triangles) and JIA (circles) with new diagnosis JIA denoted (red circle). (B) IL-17 produced by Th1 and Th17 cells for HC^C^ (triangles) and JIA (circles) with new diagnosis JIA denoted (red circle). IL-17 was below detection for Th2 cells. (C) TNFα produced by Th1 and Th17 cells for HC^C^ (triangles) and JIA (circles) with new diagnosis JIA denoted (red circle). TNFα was below detection for Th2 cells. JIA (N=19) and child control (N=20). Each Th cell culture was performed in technical triplicate and the average used for analysis. Shown is mean with standard deviation. * P < 0.05 by Mann-Whitney test.

To determine if differences in cytokine production were due T cell subset frequencies, HC^C^ and JIA Th1, Th2, and Th17 cells were analyzed by flow cytometry, and no differences were found in the frequencies of CD3^+^CD4^+^ and CD3^+^CD8^+^ cells in Th cultures (Supplemental Figure 3).

### Increased expression of mRNAs encoding IFNγ, IL-17, T-bet and RORγT in JIA Th1 cells

Th differentiation requires STAT signaling, master transcription factor induction, and signature cytokine expression. We assessed these genes expression levels in JIA and HC^C^ Th1 and Th2 culture cells by quantitative real time qPCR. The inflammatory cytokines IFNγ and IL17 were expressed higher in Th1 than Th2 cultures, with IFNγ expression magnitudes higher than IL-17 (Figure 3A). Importantly, IFNγ and IL-17 were expressed at significantly higher levels in JIA Th1 cultures than in HC^C^ (Figure 3A). Master transcription factors Tbet, RORγT, and GATA3 are required for Th1, Th17, and Th2 lineage commitment, respectively. We found Tbet and RORγT were more highly expressed in Th1 cultures and GATA3 was more highly expressed in Th2 cultures (Figure 3B). Importantly, both Tbet and RORγT were expressed significantly higher in JIA Th1 cultures than HC^C^ (Figure 3B). In STAT signaling, STAT4 and STAT1 increase Tbet and are important in Th1 differentiation and STAT3 increases RORγT and is important in Th17 differentiation. STAT4, STAT1, and STAT3 gene expression levels were not different between JIA and HC^C^ in Th1 and Th2 cultures (Supplemental Figure 4).

**Fig. 3.**
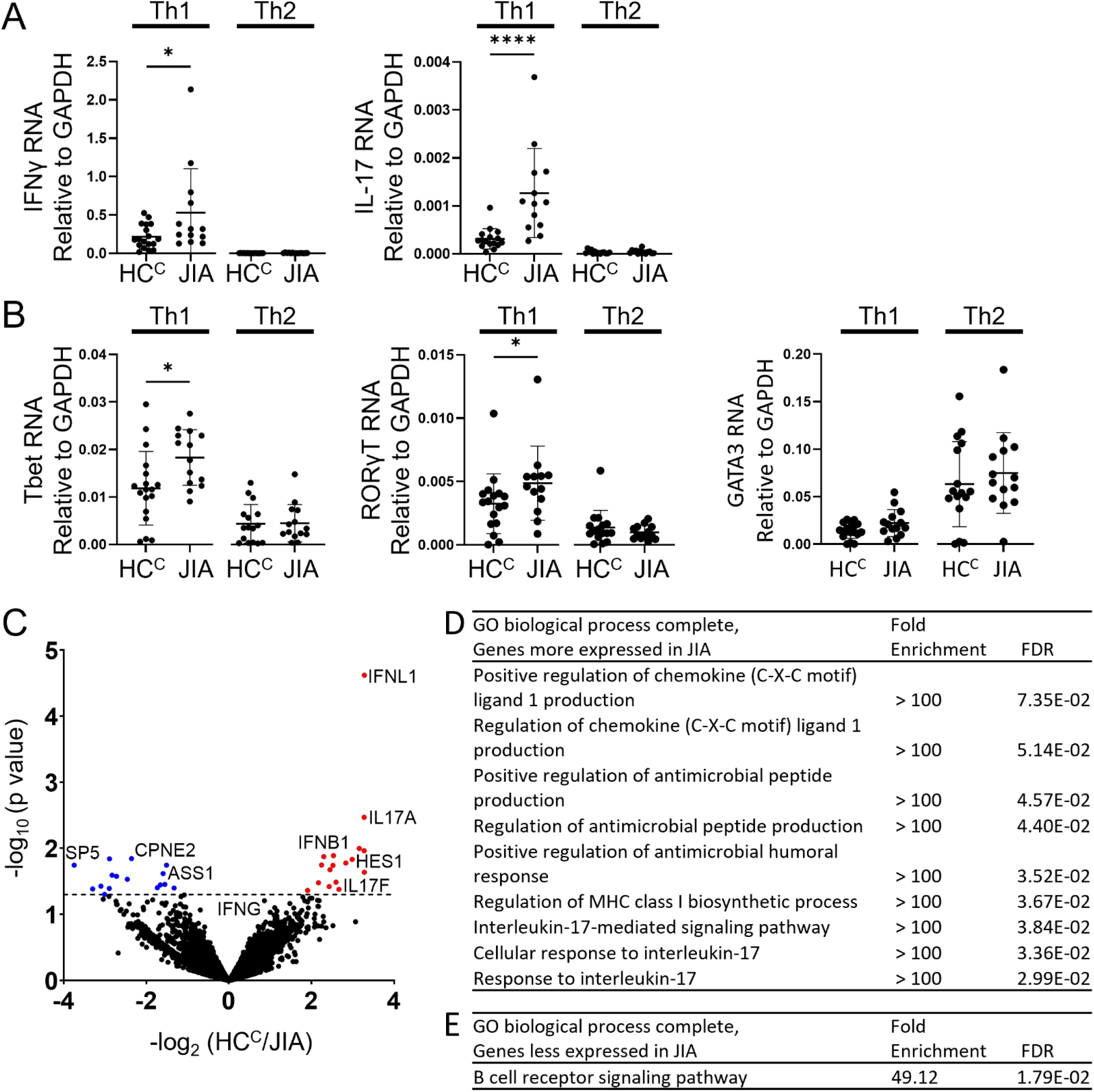
Altered gene expression and Th1 biologic pathway activation in JIA. Gene expression was examined in JIA Th1 and Th2 cultures by RT-PCR and RNA sequencing. (A) IFNγ and IL-17 RNA expression in HC^C^ and JIA Th1 and Th2 cells using quantitative RT-PCR. B) Tbet, RORγT, and GATA3 RNA expression in HC^C^ and JIA Th1 and Th2 cells using quantitative RT-PCR. For RT-PCR, JIA Th1 (N=14), HC^C^ Th1 (N=18), JIA Th2 (N=14) and HC^C^ Th2 (N=16) with gene expression relative to GAPDH and definite outliers removed. Shown is mean and standard deviation. * p<0.05, **** P<0.0001 by Mann-Whitney test. (C) Gene expression in Th1 cells from 3 child healthy controls (HC^C^) and 3 JIAs was analyzed with RNA sequencing. Differentially expressed genes assessed by DESeq2. Volcano plot of differentially expressed genes between HC^C^ and JIA with genes with p value <0.05 shown with higher expression in JIA (red circles) and lower expression in JIA (blue circles). (D) PANTHER Overrepresentation Test from the GO Ontology database for genes overexpressed in JIA. (E) PANTHER Overrepresentation Test from the GO Ontology database for genes underexpressed in JIA.

Biological pathway differences in JIA and HC^C^ Th1 cells were determined by RNA sequencing and pathways analysis. RNA from 3 HC^C^ and 3 JIA Th1 cultures were analyzed for differential gene expression using DeSeq2. Genes with p-value less than 0.05 were identified, finding 17 genes with increased and 16 genes with decreased expression in JIA (Figure 3C) (Supplemental Table 1). The most overexpressed JIA Th1 protein coding genes were IFNL1 and IL17A. The JIA genes with increased expression were analyzed for enriched biological processes using a PANTHER overrepresentation test. The identified biologic processes included important immune pathways and the IL-17 mediated signaling pathway (Figure 3D). The JIA genes with decreased expression were analyzed for enriched biologic processes, finding enrichment in the B cell receptor signaling pathway (Figure 3E). Thus, RNA sequencing data identified pathways associated with IL-17 as highly dysregulated in JIA Th1 cells.

### Elevated IFNγ and IL-17 production in a subset of JIA patients

We compared IFNγ and IL-17 production levels in individual samples to determine if heightened IFNγ and IL-17 production is similar across the studied JIA patients or specific to a group of these JIA patients. We compared ELISA data from Th1 and Th17 cell cultures from all 39 patient PBMCs. The JIA and HC^C^ Th1 cell IFNγ production was ranked from low to high, and 13 JIAs were in the highest 20 IFNγ producers (Figure 4A). The JIA and HC^C^ Th1 cell IL-17 production was ranked from low to high, and 13 JIA cultures were in the highest 20 IL-17 producers (Figure 4B). The JIA and HC^C^ Th17 cell IL-17 production was ranked from low to high, and 14 JIAs were in the highest 20 IL-17 producers (Figure 4C). In all 3 sets, JIA was significantly higher than HC^C^ by Mann-Whitney test, and the highest 20 cytokine producers included more JIA than HC^C^. The highest 20 cytokine producers from each set were combined and included 32 unique patient PBMCs. The patient PBMCs present in more than one set were identified. We found 10 patient PBMCs were present in all 3 sets, which included 9 JIAs (JIA^IFNγ-IL17 Overlap^) and 1 HC^C^ (Figure 4D). We compared the clinical characteristics of the JIA^IFNγ-IL17 Overlap^ to the remaining JIA patients (JIA^Other^) and found no differences in physician global assessment score, active joints, and treatment with biologic (Supplemental Table 2). We identified that a smaller group of the studied JIA patients more strongly exhibits the high IFNγ and IL-17 phenotype.

**Figure 4.**
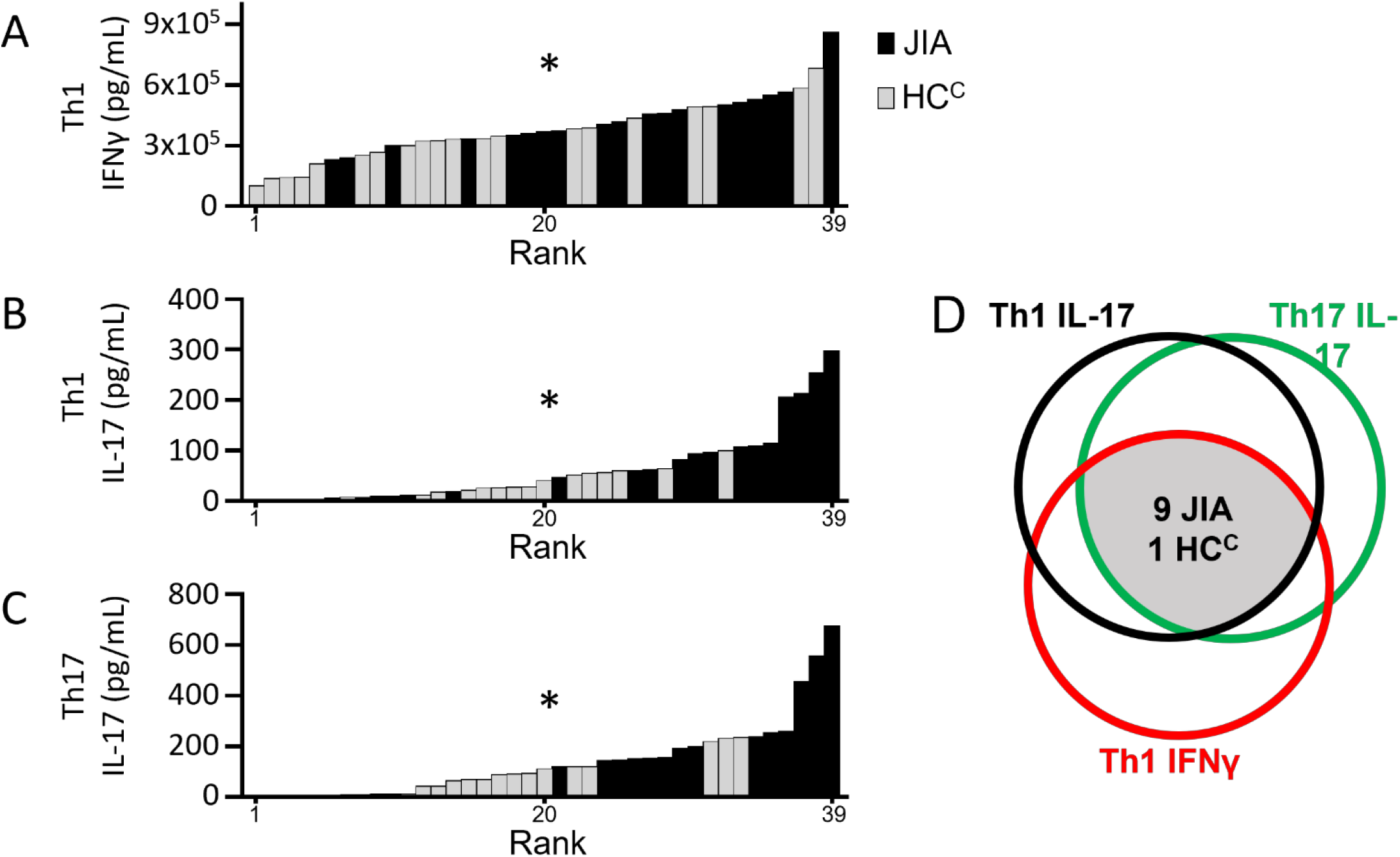
A Comparison of JIA Th1 and Th17 IFNγ and IL-17 production. Th1 and Th17 cell production of IFNγ and IL-17 measured by ELISA were ranked from low to high and compared to identify overlap in high cytokine producing patients. (A) Th1 cell IFNγ production, (B) Th1 cell IL-17 production, and (C) Th17 cell culture IL-17 production with patient PBMCs ranked from low to high with child control (HC^C^)(N=20, light gray) and JIA (N=19, black). Comparison of HC^C^ and JIA by Mann-Whitney test P < 0.05. (D) The top 20 cytokine producers in each group were compared to identify overlapping samples. A Venn diagram depicts sample overlap with 9 JIA and 1 HC^C^ present in all 3 groups.

### Increased JIA Th1 dual IFNγ and IL-17 producing cells

We next asked if increased IFNγ and IL-17 JIA Th1 cultures were derived from single or dual cytokine producing cells. JIA Th1 cultures from 8 patients were analyzed by flow cytometry for frequency of cells producing IFNγ, IL-17 or both IFNγ and IL-17. A representative JIA patient Th1 culture flow cytometry diagram showed cells producing IFNγ, IL-17, and both IFNγ and IL-17 (Figure 5A). Frequencies of IFNγ^+^IL-17^-^, IFNγ^-^IL-17^-^, IFNγ^-^IL-17^+^, and IFNγ^+^IL-17^+^ cells were determined. We found single IFNγ producing cells were the highest frequency cytokine producing cells in the Th1 culture (Figure 5B). We also found frequencies of IL-17 producing cells that could be sub-divided into IL-17 single producers and IL-17^+^IFNγ^+^ dual producers. We then determined the frequency of Tbet expressing cells in the IFNγ^+^IL-17^-^, IFNγ^-^IL-17^-^, IFNγ^-^IL-17^+^, and IFNγ^+^IL-17^+^ cell subsets. We found that Tbet frequency was highest in IFNγ^+^IL-17^+^ cells, which was significantly higher than the IFNγ^-^IL-17^-^ cells (Figure 5C). In summary, we found that JIA Th1 polarization from peripheral blood generated cells producing IL-17 and dual IFNγ-IL-17 producing cells.

**Figure 5.**
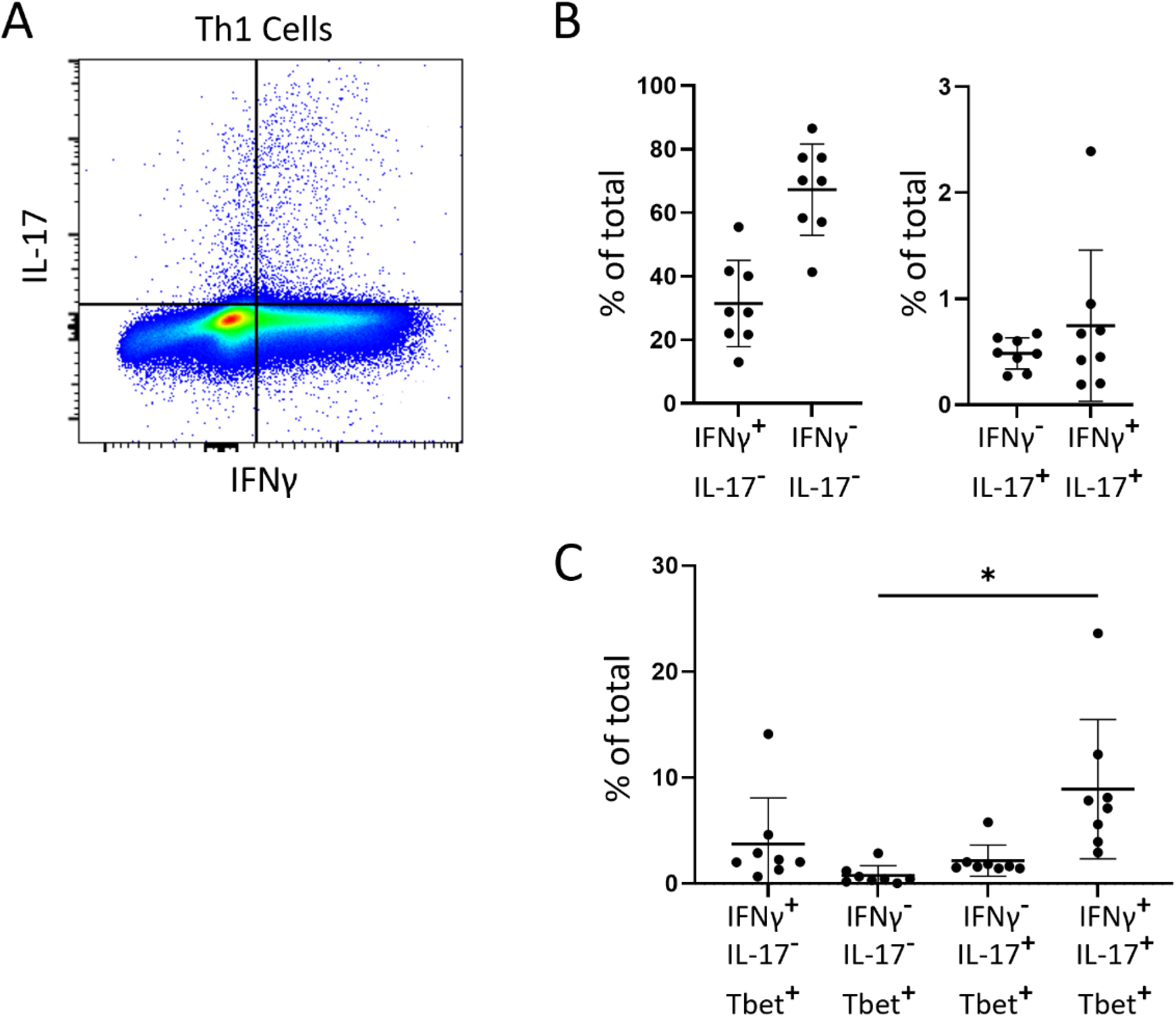
JIA Th1 cells have dual IFNγ^+^IL-17^+^ cells expressing Tbet. JIA Th1 culture cells were assessed with flow cytometry to identify IFNγ^+^ and IL-17^+^ cell subsets. Fluorescent minus one controls were used to identify positive populations. (A) Representative flow cytometry plot from a JIA patient Th1 cells showing IFNγ and IL-17 positivity and gating. (B) Analysis of JIA Th1 culture cells showing frequency of IFNγ and IL-17 positivity per live cells (N=8). IL-17 negative and positive cells are shown on different graphs. (C) Analysis of JIA Th1 culture cells showing frequency of Tbet positivity in the IFNγ^+^IL-17^-^, IFNγ^-^IL-17^-^, IFNγ^-^IL-17^+^, and IFNγ^+^IL-17^+^ cell subsets. Shown is mean with standard deviation. * p < 0.05 by ANOVA test.

## Discussion

In this study, we demonstrate that T cells from JIA patients develop an inflammatory cytokine profile in response to Th1 and Th17 differentiation stimuli. On initial assessment, JIA peripheral T cells are similar to HC^C^ with the same naïve and memory T cell profiles and proliferative capacity. After Th1 differentiation, JIA cells express increased IFNγ and IL-17 and increased IFNγ, IL-17, Tbet and RORγT mRNA. This is surprising since Th1 cultures should effectively suppress the Th17 differentiation that is associated with RORγT and IL-17. This contrasts with Th1 differentiation in HC^C^ cells where both RORγT and IL-17 are suppressed. JIA Th1 differentiation leads to single IFNγ, single IL-17, and dual IFNγ-IL-17 producing cells. The dual IFNγ-IL-17 producing cells are similar to previously described Th1.17 cells. Under Th17 polarizing conditions, JIA T cells produce increased IL-17. Under Th2 differentiation conditions, JIA T cells do not express IFNγ or IL-17. Our work demonstrates that peripheral T cells in JIA develop a pro-inflammatory profile using a well-established Th differentiation protocol. Using this assay, studies to reveal the genetic, epigenetic, and environmental factors contributing to the enhanced IFNγ and IL-17 phenotype and Th1.17 cell development can be pursued.

A striking finding is the similarity between JIA and HC^C^ peripheral T cells. Similarly, polyarticular RF-JIA patients and controls do not exhibit different memory and naïve T cell phenotypes (13). Despite the similarities in peripheral lymphocytes, a subset of hyperresponsive JIA CD4^+^ T cells did respond more strongly to IFNγ (13). Additionally, JIA and HC^C^ T cell proliferative capacity is similar. A limitation in this study is that a small population of dividing cells or an antigen specific population may not be identified. We also did not study the same individuals at different time points; thus, cells could vary with time. Importantly, we minimize age dependent immune effects by focusing on a prepubescent population in both JIA and HC^C^.

JIA synovial fluid has an increase in Th1 cells, Th17 cells, and associated cytokines for oligoarticular, polyarticular, and enthesitis-arthritis subtypes (18-20, 30, 31). This parallels our Th culture findings that Th1 and Th17 differentiation produce increases in IFNγ and IL-17, however our cultures are from peripheral blood. An unexpected result from our Th1 cultures is the increase in IL-17. Our JIA Th17 cultures produce more IL-17 and suppress IFNγ.

Dual IFNγ and IL-17 producing Th cells, called Th17.1, are identified in various autoimmune conditions, often at sites of inflammation (33-35). JIA synovial fluid contains Th17.1 cells (22, 35). Th17.1 cells begin as Th17 cells that produce IFNγ in response to IL-12, then dual producing cells stop producing IL-17 and become sole IFNγ producers (22). To our knowledge, no studies generate Th17.1 cells from JIA patient peripheral T cells. In our study, we call the JIA dual IFNγ and IL-17 producing cells Th1.17 cells. The JIA Th1 cultures have more IFNγ, IL-17, Tbet and RORγT at the gene expression level. Normally, IL-12 drives Th1 differentiation that increases Tbet and IFNγ and suppresses RORγT and IL-17 expression (36). IL-12 also shifts the Th17.1 cells to become sole IFNγ producers (22). Our culture conditions result in different findings, with IL-12 driving generation of dual-producing cells and RORγT and IL-17 expression. How Th1.17 cells develop during Th1 differentiation is unknown. One hypothesis is that JIA cells are resistant to IFNγ-induced suppression of Th17 pathways, resulting in IFNγ and IL-17 dual producers. Another hypothesis is that JIA naïve T cells have an abnormal response to IL-12 resulting in dual activation of Tbet and RORγT driven pathways. Additional studies are necessary to differentiate between these hypotheses. Importantly, the JIA inflammatory Th1 and Th17 phenotype is suppressed by Th2 differentiation suggesting that counterregulatory pathways could prevent inflammatory cell development.

Biologic heterogeneity is present in JIA, and biologic phenotypes do not always clearly align with clinical phenotypes (6). We identified a subset of JIA PBMCs with a biological phenotype of high IFNγ and IL-17. In our study the IFNγ and IL-17 phenotype did not correlate with differences in physician global assessment scores, active joints, or exposure to biologic medications. This suggests the biologic phenotype arises in response to other sample or patient characteristics in a subset of JIA patients.

How heterogeneity in JIA develops and whether it is due to genetics or environment or a combination of both is poorly understood. For example, we identified a JIA patient with a monogenetic loss of function mutation in GATA3 that exhibits a similar phenotype to the patients herein, with increased IFNγ and IL17 in Th1 cultures and increased IL17 in Th17 cultures (24). The molecular basis of this finding is due to loss of GATA3 function. Another study has identified rare variants and their association with immune pathways in JIA (37). An important question is, do rare mutations in common pathways contribute to JIA development of IFNγ and IL-17 producing cells? Determining whether JIA T cells have an innate tendency to become IFNγ and IL-17 producing cells, and how genetic mutations might contribute to this phenotype, may open new avenues for understanding disease onset pathogenesis and developing personalized approaches to JIA medication choices and diagnostics.

## Data Availability

All data included in this study are available upon request by contacting the corresponding author. The sequences presented in this article have been submitted to the National Center for Biotechnology Information Gene Expression Omnibus (https://www.ncbi.nlm.nih.gov/geo/).

## Acknowledgements

The authors thank the patients, families, and clinical staff for their contributions.

## Ethics

This study complies with the Declaration of Helsinki, the locally appointed ethics committee has approved the research protocol and that informed consent has been obtained from the subjects (or their legally authorized representative).

## References

1. Oberle EJ, Harris JG, Verbsky JW. Polyarticular juvenile idiopathic arthritis - epidemiology and management approaches. Clin Epidemiol. 2014;6:379–93.

2. Hinze C, Gohar F, Foell D. Management of juvenile idiopathic arthritis: hitting the target. Nat Rev Rheumatol. 2015;11(5):290–300.

3. Guzman J, Oen K, Huber AM, Watanabe Duffy K, Boire G, Shiff N, et al. The risk and nature of flares in juvenile idiopathic arthritis: results from the ReACCh-Out cohort. Ann Rheum Dis. 2016;75(6):1092–8.

4. Guzman J, Oen K, Tucker LB, Huber AM, Shiff N, Boire G, et al. The outcomes of juvenile idiopathic arthritis in children managed with contemporary treatments: results from the ReACCh-Out cohort. Ann Rheum Dis. 2015;74(10):1854–60.

5. Brunner HI, Schanberg LE, Kimura Y, Dennos A, Co DO, Colbert RA, et al. New Medications Are Needed for Children With Juvenile Idiopathic Arthritis. Arthritis Rheumatol. 2020;72(11):1945–51.

6. Eng SW, Duong TT, Rosenberg AM, Morris Q, Yeung RS, Reacch OUT, et al. The biologic basis of clinical heterogeneity in juvenile idiopathic arthritis. Arthritis Rheumatol. 2014;66(12):3463–75.

7. Petty RE, Southwood TR, Baum J, Bhettay E, Glass DN, Manners P, et al. Revision of the proposed classification criteria for juvenile idiopathic arthritis: Durban, 1997. J Rheumatol. 1998;25(10):1991–4.

8. Sullivan DB, Cassidy JT, Petty RE. Pathogenic implications of age of onset in juvenile rheumatoid arthritis. Arthritis Rheum. 1975;18(3):251–5.

9. van den Broek T, Borghans JAM, van Wijk F. The full spectrum of human naive T cells. Nat Rev Immunol. 2018;18(6):363–73.

10. Saule P, Trauet J, Dutriez V, Lekeux V, Dessaint JP, Labalette M. Accumulation of memory T cells from childhood to old age: central and effector memory cells in CD4(+) versus effector memory and terminally differentiated memory cells in CD8(+) compartment. Mech Ageing Dev. 2006;127(3):274–81.

11. Prelog M, Schwarzenbrunner N, Tengg E, Sailer-Hock M, Kern H, Zimmerhackl LB, et al. Quantitative alterations of CD8+ T cells in juvenile idiopathic arthritis patients in remission. Clin Rheumatol. 2009;28(4):385–9.

12. Prelog M, Schwarzenbrunner N, Sailer-Hock M, Kern H, Klein-Franke A, Ausserlechner MJ, et al. Premature aging of the immune system in children with juvenile idiopathic arthritis. Arthritis and rheumatism. 2008;58(7):2153–62.

13. Throm AA, Moncrieffe H, Orandi AB, Pingel JT, Geurs TL, Miller HL, et al. Identification of enhanced IFN-gamma signaling in polyarticular juvenile idiopathic arthritis with mass cytometry. JCI Insight. 2018;3(15).

14. Zhu J, Yamane H, Paul WE. Differentiation of effector CD4 T cell populations (*). Annu Rev Immunol. 2010;28:445–89.

15. Schraml BU, Hildner K, Ise W, Lee WL, Smith WA, Solomon B, et al. The AP-1 transcription factor Batf controls T(H)17 differentiation. Nature. 2009;460(7253):405–9.

16. Zhu J, Paul WE. Peripheral CD4+ T-cell differentiation regulated by networks of cytokines and transcription factors. Immunol Rev. 2010;238(1):247–62.

17. Raphael I, Nalawade S, Eagar TN, Forsthuber TG. T cell subsets and their signature cytokines in autoimmune and inflammatory diseases. Cytokine. 2015;74(1):5–17.

18. Nistala K, Moncrieffe H, Newton KR, Varsani H, Hunter P, Wedderburn LR. Interleukin-17-producing T cells are enriched in the joints of children with arthritis, but have a reciprocal relationship to regulatory T cell numbers. Arthritis Rheum. 2008;58(3):875–87.

19. Wedderburn LR, Robinson N, Patel A, Varsani H, Woo P. Selective recruitment of polarized T cells expressing CCR5 and CXCR3 to the inflamed joints of children with juvenile idiopathic arthritis. Arthritis Rheum. 2000;43(4):765–74.

20. Mahendra A, Misra R, Aggarwal A. Th1 and Th17 Predominance in the Enthesitis-related Arthritis Form of Juvenile Idiopathic Arthritis. J Rheumatol. 2009;36(8):1730–6.

21. Maggi L, Cosmi L, Simonini G, Annunziato F, Cimaz R. T cell subpopulations in juvenile idiopathic arthritis and their modifications after biotherapies. Autoimmun Rev. 2016;15(12):1141–4.

22. Nistala K, Adams S, Cambrook H, Ursu S, Olivito B, de Jager W, et al. Th17 plasticity in human autoimmune arthritis is driven by the inflammatory environment. Proc Natl Acad Sci U S A. 2010;107(33):14751–6.

23. Spurlock CF, 3rd, Tossberg JT, Guo Y, Collier SP, Crooke PS, 3rd, Aune TM. Expression and functions of long noncoding RNAs during human T helper cell differentiation. Nat Commun. 2015;6:6932.

24. Patrick AE, Wang W, Brokamp E, Graham TB, Aune TM, Duis JB. Juvenile idiopathic arthritis associated with a mutation in GATA3. Arthritis Res Ther. 2019;21(1):156.

25. Petty RE, Southwood TR, Manners P, Baum J, Glass DN, Goldenberg J, et al. International League of Associations for Rheumatology classification of juvenile idiopathic arthritis: second revision, Edmonton, 2001. J Rheumatol. 2004;31(2):390–2.

26. Love MI, Huber W, Anders S. Moderated estimation of fold change and dispersion for RNA-seq data with DESeq2. Genome Biol. 2014;15(12):550.

27. Ashburner M, Ball CA, Blake JA, Botstein D, Butler H, Cherry JM, et al. Gene ontology: tool for the unification of biology. The Gene Ontology Consortium. Nat Genet. 2000;25(1):25–9.

28. Gene Ontology C. The Gene Ontology resource: enriching a GOld mine. Nucleic Acids Res. 2021;49(D1):D325–D34.

29. Mi HY, Muruganujan A, Ebert D, Huang XS, Thomas PD. PANTHER version 14: more genomes, a new PANTHER GO-slim and improvements in enrichment analysis tools. Nucleic Acids Research. 2019;47(D1):D419–D26.

30. Jule AM, Hoyt KJ, Wei K, Gutierrez-Arcelus M, Taylor ML, Ng J, et al. Th1 polarization defines the synovial fluid T cell compartment in oligoarticular juvenile idiopathic arthritis. JCI Insight. 2021.

31. Aggarwal A, Agarwal S, Misra R. Chemokine and chemokine receptor analysis reveals elevated interferon-inducible protein-10 (IP)-10/CXCL10 levels and increased number of CCR5+ and CXCR3+ CD4 T cells in synovial fluid of patients with enthesitis-related arthritis (ERA). Clin Exp Immunol. 2007;148(3):515–9.

32. Omoyinmi E, Hamaoui R, Pesenacker A, Nistala K, Moncrieffe H, Ursu S, et al. Th1 and Th17 cell subpopulations are enriched in the peripheral blood of patients with systemic juvenile idiopathic arthritis. Rheumatology (Oxford). 2012;51(10):1881–6.

33. Annunziato F, Cosmi L, Santarlasci V, Maggi L, Liotta F, Mazzinghi B, et al. Phenotypic and functional features of human Th17 cells. J Exp Med. 2007;204(8):1849–61.

34. Stadhouders R, Lubberts E, Hendriks RW. A cellular and molecular view of T helper 17 cell plasticity in autoimmunity. J Autoimmun. 2018;87:1–15.

35. Cosmi L, Cimaz R, Maggi L, Santarlasci V, Capone M, Borriello F, et al. Evidence of the transient nature of the Th17 phenotype of CD4+CD161+ T cells in the synovial fluid of patients with juvenile idiopathic arthritis. Arthritis Rheum. 2011;63(8):2504–15.

36. McGeachy MJ, Cua DJ. Th17 cell differentiation: the long and winding road. Immunity. 2008;28(4):445–53.

37. Meng X, Hou X, Wang P, Glessner JT, Qu HQ, March ME, et al. Association of novel rare coding variants with juvenile idiopathic arthritis. Ann Rheum Dis. 2021;80(5):626–31.

