## Supplemental Data for "JIA patient T cells differentiate into Th1, Th17 and Th1.17 effector cells under Th1 polarizing conditions"

**Supplemental Fig. 1 T cell subsets from adult and child healthy controls and JIA are the same.**

Adult healthy control ($HC^A$), child healthy control ($HC^C$), and JIA PBMCs were analyzed with flow cytometry for T cell subsets of $CD3^+$, $CD4^+$, and $CD8^+$ positive cells. (A) Live $CD3^+$ cell representative flow cytometry plot. (B) Analysis of the frequency of live $CD3^+$ cells. (C) Analysis of the frequency of $CD3^+CD4^+$ and $CD3^+CD8^+$ cells per total live cells. $HC^A$ (N=4), $HC^C$ (N=20), JIA (N=20). Shown is mean with standard deviation. Analysis by Mann-Whitney test with * $p<0.05$. No significant differences.

**Supplemental Fig. 2 T cell proliferation from child control and JIA are the same.**

A T cell proliferation assay was performed by staining child healthy control ($HC^C$) and JIA PBMCs with the fluorophore CFSE, stimulating cells with anti-CD3 and anti-CD28, and on day 3 measuring T cell subset proliferation using flow cytometry analysis for $CD3^+$, $CD4^+$, and $CD8^+$ cells. A proliferation index (PI) was calculated by measuring the number of divisions per dividing cell. (A) Representative flow cytometry plot for CFSE proliferation of $CD3^+$ cells. (B) Analysis of PIs for $CD3^+$, $CD3^+CD4^+$, and $CD3^+CD8^+$ cells for $HC^C$ (N=14) and JIA (N=11). (C) The T cell proliferation assay was preformed in the presence of IL-12 to drive Th1 polarization. Analysis of PIs for $CD3^+$, $CD3^+CD4^+$, and $CD3^+CD8^+$ cells for $HC^C$ (N=5) and JIA (N=3). (D) The T cell proliferation assay for $CD3^+$ cells without (N=25) and with Th1 polarization by IL-12 (N=8) were compared by combining pediatric samples of $HC^C$ (black circle) and JIA (empty circle). Shown is mean with standard deviation. Analysis by Mann-Whitney test with * $p<0.05$, ** $p<0.01$.

**Supplemental Fig. 3 T cell subsets in Th1, Th2, and Th17 cell cultures from child control and JIA.**

Th1, Th2, and Th17 cell cultures were prepared from child healthy control ($HC^C$) and JIA. The polarized cells were analyzed by flow cytometry for $CD3^+CD4^+$ and $CD3^+CD8^+$ T cell subsets. (A) Analysis of live $CD3^+CD4^+$ and $CD3^+CD8^+$ cell frequencies in Th1 cell cultures from $HC^C$ (N=11) and JIA (N=11). (B) Analysis of live $CD3^+CD4^+$ and $CD3^+CD8^+$ cell frequencies in Th2 cell cultures from $HC^C$ (N=11) and JIA (N=10). (C) Analysis of live $CD3^+CD4^+$ and $CD3^+CD8^+$ cell frequencies in Th17 cell cultures from $HC^C$ (N=11) and JIA (N=10). Shown is mean with standard deviation. Analysis by Mann-Whitney test with * $p<0.05$. No significant differences.

**Supplemental Fig. 4 STAT gene expression in healthy control and JIA Th cells.**

Gene expression of the STAT transcription factors STAT4, STAT1, and STAT3 were examined in Th1 and Th2 cultures from child healthy controls ($HC^C$) and JIAs using quantitative RT-PCR. (A) STAT4, STAT1, and STAT3 RNA expression in $HC^C$ and JIA Th1 and Th2 cells. JIA Th1 (N=14), $HC^C$ Th1 (N=18), JIA Th2 (N=14) and $HC^C$ Th2 (N=16). Gene expression is relative to GAPDH with definite outliers removed. Shown is mean and standard deviation. Analysis by Mann-Whitney test with * $p<0.05$. No significant differences.

**Supplemental Table 1. Th1 cell differentially expressed genes between JIA and child control with p <0.05.**

**Supplemental Table 2. Characteristics of JIA with overlapping high IFNγ and IL-17 in Th1 and Th17 cells and other JIA.**

**Supplemental Fig. 1**

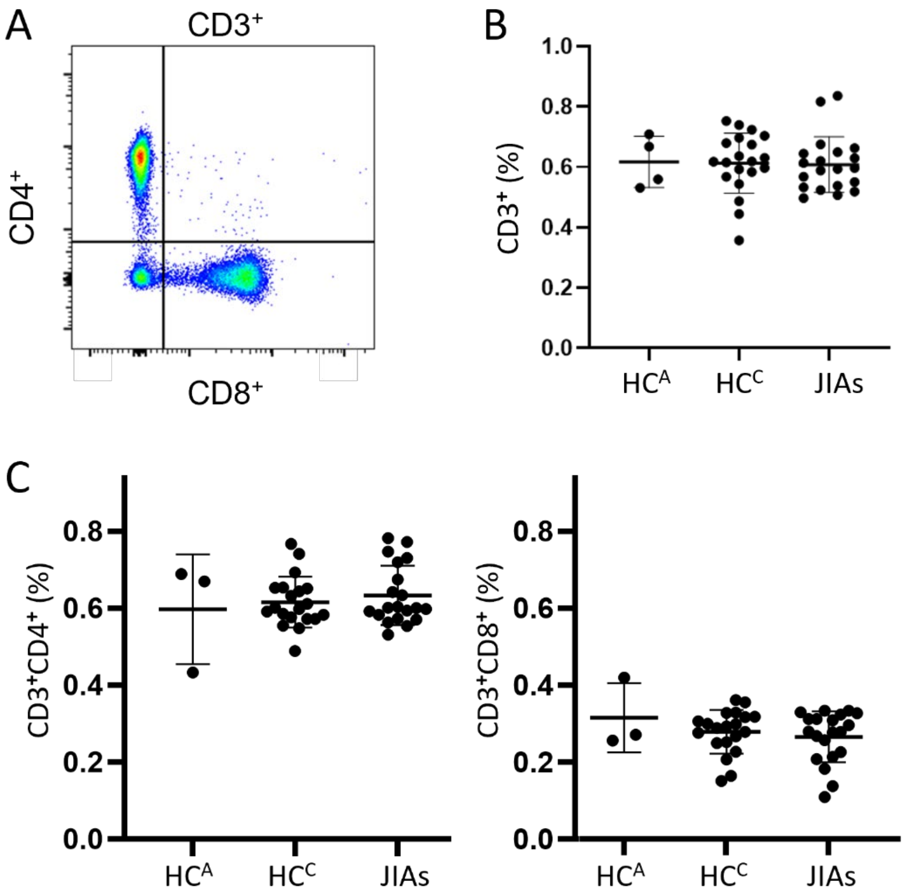

**Supplemental Fig. 2**

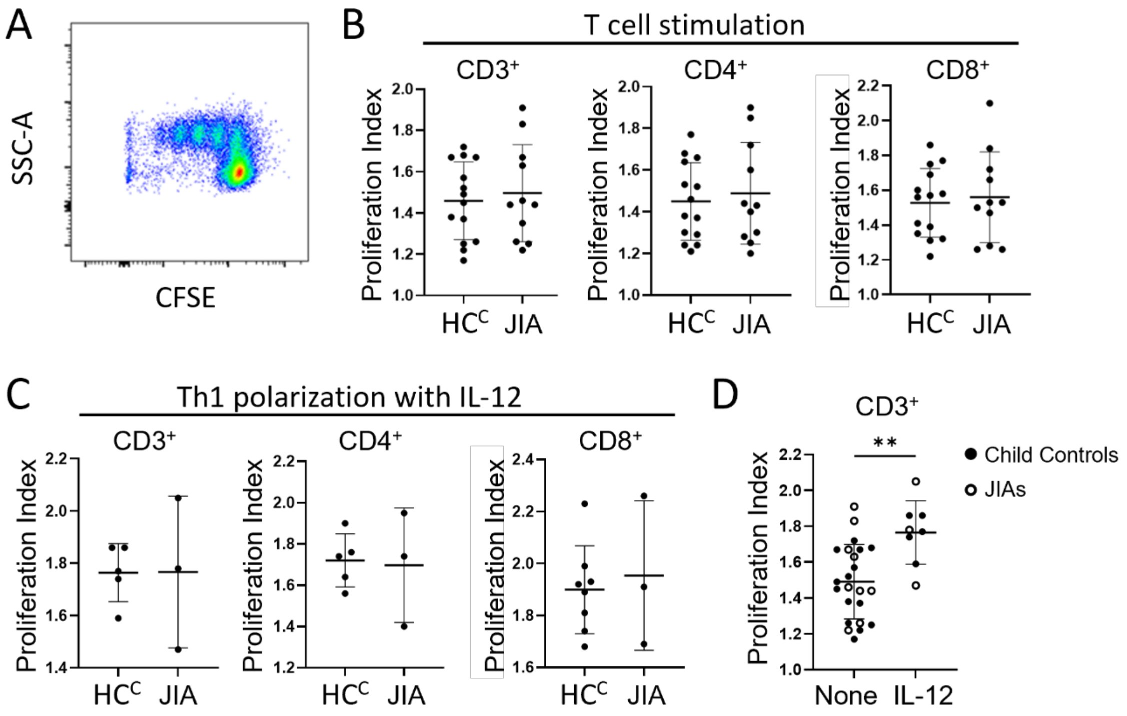

**Supplemental Fig. 3**

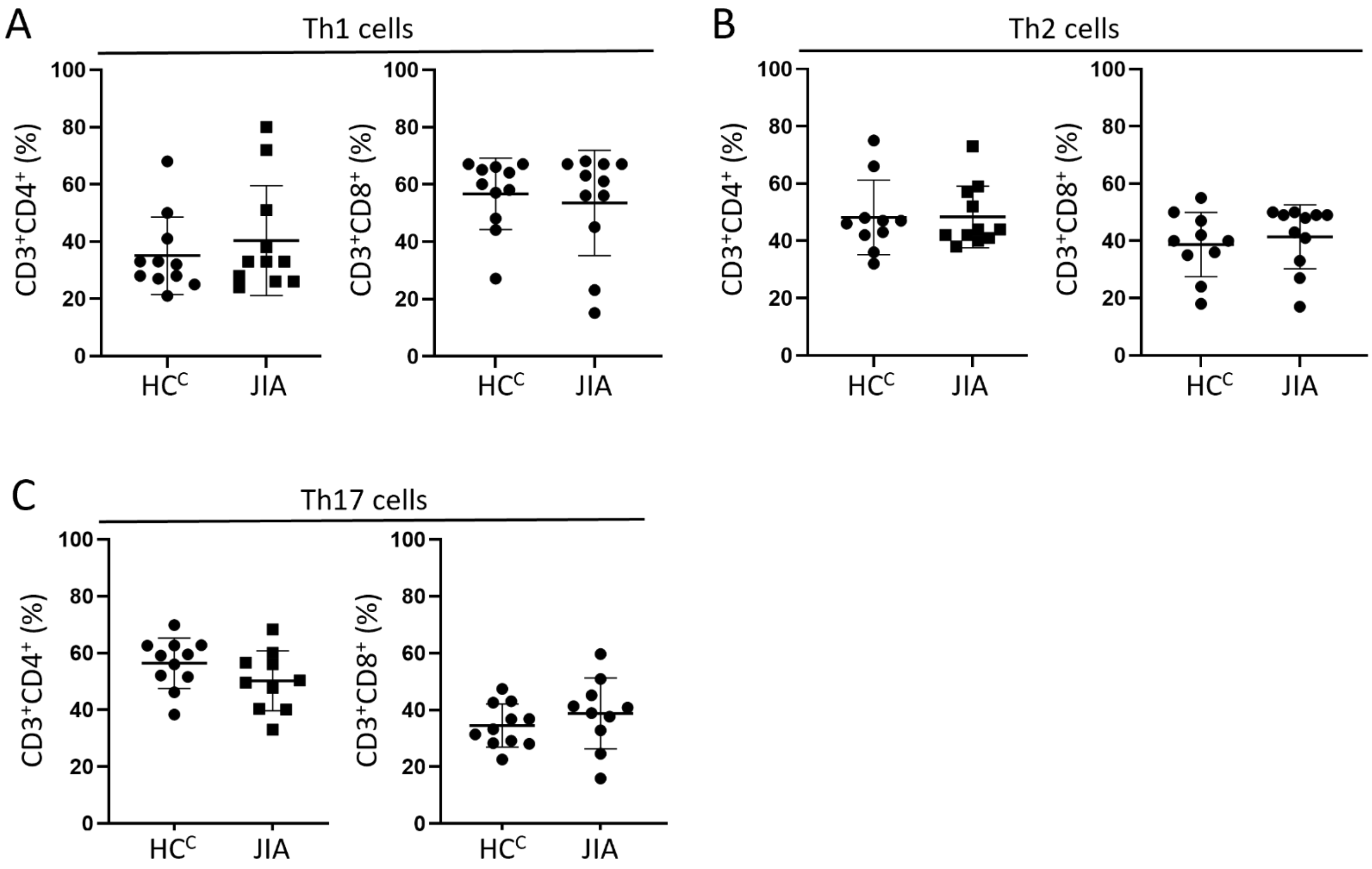

**Supplemental Fig. 4**

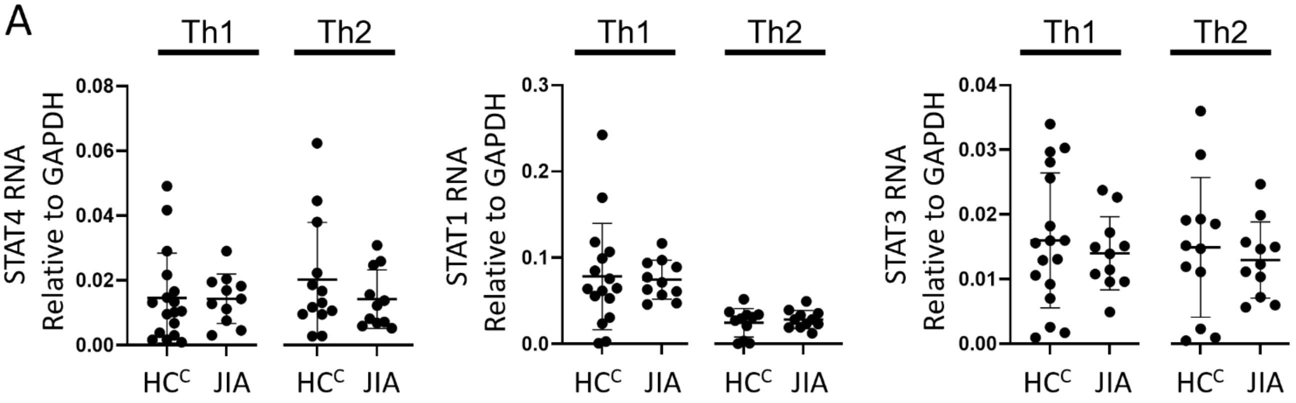

### Supplemental Table 1

| | baseMean | log2FoldChange | lfcSE | stat | pvalue | Ensemble |
|---|---|---|---|---|---|---|
| LINC00887 | 229.5036228 | -3.282064262 | 1.444277997 | -2.272460198 | 0.023058729 | ENSG00000214145 |
| IFNL1 | 258.1045835 | -3.280737223 | 0.776968794 | -4.222482611 | 2.42E-05 | ENSG00000182393 |
| IL17A | 270.8269026 | -3.277124939 | 1.119003182 | -2.928610921 | 0.003404803 | ENSG00000112115 |
| TSKS | 235.5305558 | -3.275720858 | 1.286906837 | -2.545421908 | 0.010914577 | ENSG00000126467 |
| CTD.3222D19.12 | 294.3359079 | -3.158557878 | 1.227074285 | -2.574055962 | 0.010051401 | ENSG00000269399 |
| HES1 | 126.7289088 | -2.987802108 | 1.224955559 | -2.439110617 | 0.014723461 | ENSG00000114315 |
| RP5.890E16.2 | 229.4133056 | -2.833468025 | 1.184252234 | -2.392622066 | 0.016728463 | ENSG00000263412 |
| IL17F | 2317.845574 | -2.66606902 | 1.309882749 | -2.035349364 | 0.041815732 | ENSG00000112116 |
| TMEM213 | 189.6759105 | -2.595901796 | 1.214562309 | -2.137314634 | 0.032572409 | ENSG00000214128 |
| IFNB1 | 161.8574342 | -2.532044582 | 1.018380609 | -2.486344065 | 0.012906312 | ENSG00000171855 |
| SPSB2 | 512.4596369 | -2.51991147 | 1.066840919 | -2.362031139 | 0.018175116 | ENSG00000111671 |
| ALDH4A1 | 317.4531246 | -2.452131272 | 1.063467843 | -2.305787889 | 0.021122487 | ENSG00000159423 |
| PLAT | 264.691779 | -2.428523342 | 1.169882525 | -2.075869406 | 0.037906031 | ENSG00000104368 |
| RP4.575N6.4 | 314.7985737 | -2.298054783 | 0.928821924 | -2.474160787 | 0.013354958 | ENSG00000225938 |
| CBLN3 | 284.0859812 | -2.244566683 | 0.947984473 | -2.367725155 | 0.017897826 | ENSG00000139899 |
| OLIG3 | 235.0267266 | -2.170276142 | 1.019424638 | -2.128922592 | 0.033260665 | ENSG00000177468 |
| LGALS17A | 121.6447874 | -1.906670276 | 0.94451033 | -2.018686524 | 0.043519812 | ENSG00000226025 |
| RPL23AP7 | 681.8760017 | 1.326339363 | 0.645494049 | 2.054766215 | 0.039901591 | ENSG00000240356 |
| IGFBP4 | 1703.92069 | 1.50895516 | 0.638139996 | 2.364614611 | 0.018048842 | ENSG00000141753 |
| CD79A | 1196.922598 | 1.550079696 | 0.735851558 | 2.106511399 | 0.035159954 | ENSG00000105369 |
| ASS1 | 5514.66332 | 1.591156362 | 0.705868508 | 2.254182393 | 0.024184696 | ENSG00000130707 |
| RAB38 | 466.1593356 | 1.659537425 | 0.790881514 | 2.098338872 | 0.035875221 | ENSG00000123892 |
| PDLIM1 | 723.6751717 | 1.73225893 | 0.841364041 | 2.0588697 | 0.039506722 | ENSG00000107438 |
| CPNE2 | 545.8580394 | 2.35572233 | 0.962119066 | 2.448472764 | 0.014346328 | ENSG00000140848 |
| IGHG4 | 89.29080387 | 2.457653534 | 1.128559093 | 2.177691491 | 0.029429013 | ENSG00000211892 |
| IGFBP2 | 3038.714142 | 2.720820675 | 1.226558216 | 2.218256451 | 0.026537352 | ENSG00000115457 |
| RP11.253E3.1 | 115.599896 | 2.828023209 | 1.266370837 | 2.23317146 | 0.02553764 | ENSG00000234589 |
| IGHG1 | 1448.149994 | 2.889952643 | 1.181303481 | 2.446409995 | 0.014428682 | ENSG00000211896 |
| JCHAIN | 112.0140031 | 2.896349233 | 1.414098409 | 2.048194959 | 0.0405409 | ENSG00000132465 |
| IGHA2 | 70.74929883 | 3.009949306 | 1.535003057 | 1.960875122 | 0.049893592 | ENSG00000211890 |
| RPL23AP4 | 97.7935494 | 3.098787818 | 1.489313048 | 2.080682649 | 0.037462963 | ENSG00000212932 |
| IGHGP | 107.814981 | 3.299373358 | 1.616838141 | 2.040633057 | 0.041287314 | ENSG00000253755 |
| SP5 | 99.92999701 | 3.744806407 | 1.583113431 | 2.365469418 | 0.01800723 | ENSG00000204335 |

**Supplemental Table 2**

| | $JIA^{IFNγ\text{-}IL17\ Overlap}$ | $JIA^{Other}$ | P value* |
|---|---|---|---|
| Number of JIA | 9 | 11 | |
| Physician global assessment score, median (range) | 1 (0-5) | 1 (0-9) | 0.890 |
| Active joints, median (range) | 1 (0-10) | 0 (0-9) | 0.516 |
| Ever treatment with biologic, N (%) | 5 (55%) | 4 (40%) | |
| Current treatment with biologic, N (%) | 4 (44%) | 5 (50%) | |
